# Climate change education in public health and medical curricula in Australian and New Zealand Universities: a mixed methods study of barriers and areas for further action

**DOI:** 10.1101/2021.06.11.21258793

**Authors:** Aparna Lal, Erin Walsh, Ali Weatherell, Claudia Slimings

**Affiliations:** National Centre for Epidemiology and Population Health, Research School of Population Health, Australian National University, Canberra, ACT, Australia; Population Health Exchange, Research School of Population Health, Australian National University, Canberra, ACT, Australia; Medical School, College of Health and Medicine, Australian National University, Canberra, ACT, Australia

## Abstract

**Background:** The World Health Organization deemed climate change and air pollution as the top threat to global health in 2019. The importance of climate for health is recognised by healthcare professionals, who need to be equipped to deliver environmentally sustainable healthcare and promote planetary health. There is some evidence that climate change and health is not strongly embedded in accredited master-level public health training programs and medical programs globally, however, the immersion of climate-health in Australian and New Zealand programs is unclear.

**Objectives:** To explore the extent to which climate-health education is currently embedded into public health and medical curricula in Australia and New Zealand.

**Methods:** Educators identified by their coordination, convenorship, or delivery into programs of public health and medicine at universities in Australia and New Zealand were invited to participate in a cross-sectional, exploratory mixed methods study. Participants completed an online quantitative survey and qualitative interviews regarding their experience in program and course delivery, and the prominence of climate-health content within program and course delivery. Quantitative surveys were analysed using descriptive statistics and qualitative interview content was analysed via a modified ground theory approach.

**Results:** The response rate of the quantitative survey was 43.7% (21/48). Ten survey respondents also completed qualitative interviews. Quantitative results showed that epidemiologists were the most common experts involved in design and delivery of this curriculum, with a reliance on guest lecturers to provide updated content. Qualitative interviews highlighted the ad-hoc role of Indigenous-led content in this field, the barriers of time and resources to develop a coherent curriculum and the important role of high-level champions to drive the inclusion of climate change and planetary health.

**Conclusion:** There is an urgent need to strengthen current support available for pedagogical leadership in the area of climate and broader environmental change teaching at universities.

## Introduction

Environmental change is a major threat to global health (1). Escalating anthropogenic exploitation of the Earth’s natural resources to power economic development and population growth since the beginning of the Industrial Age has resulted in many direct and indirect hazards for human health (2, 3). These include acute, extreme weather events such as heatwaves, floods, dust storms and fires as well as slow moving, less visible environmental changes such as degradation of our land, water and air quality. In relation to human health, these changes have resulted in increased morbidity and mortality through trauma, injury and heat-related disease (4) and indirect impacts through reduced food yields, altered infectious disease patterns, loss of livelihoods, conflict, population displacement and mental health impacts (3-5). A rapidly changing natural environment will exacerbate existing socio-economic and health inequalities, with disadvantaged populations projected to experience the greatest adverse health impacts (4, 5).

Climate change and the ongoing pandemic presents us with an opportunity to redefine the environment-health curricula for the future health workforce. Health professionals play a crucial role in preventing illness that may arise from such rapid changes to our environment, through the design and implementation of sustainable healthcare interventions, health system strengthening for adaptation, promoting patient understanding and building community resilience and as advocates within their practices, communities and professional organisations (6-9). Thus, the future health workforce, including health care professionals and public health practitioners, need to be equipped with the knowledge, skills and motivation and agency to deliver sustainable healthcare and promote planetary health (10, 11). The emergence of SARS-Cov-2 virus (COVID-19) in 2019 is linked to the current environmental crisis and has brought further attention to the need for an emphasis on environmental change in health education (12).

In medical schools, there is currently a gap in formal learning and teaching opportunities – a recent survey found that only 15% of medical schools incorporated climate change and health into the curriculum in some form, and student-led activities were evident in a further 12% (13). Similarly, planetary health and climate change is not strongly embedded in accredited master-level public health training programs globally (14). There has been no focused evaluation of how these topics are embedded and taught in medical schools and public health programmes in the Southern Hemisphere. The closest research in this space is a study led by the Columbia Mailman School of Public Health that assessed climate change-health curricula internationally and included primarily medical institutions on Australia’s eastern seaboard who are members of the Global Consortium on Climate and Health Action (14, 15).

The aim of this study is to describe the extent, design and modes of delivery of planetary health and climate change educational offerings at tertiary institutions in Australia and New Zealand, including programs of both public health and medicine. In doing so, we identify some potential areas for future focus to build institutional, political and public support for more ambitious and integrated environment-health curricula.

## Methods

A cross-sectional, mixed methods study was used to assess climate-health educational offerings in Master of Public Health (MPH) and medicine degree programs at tertiary institutions in Australia and New Zealand. The sampling frame was educators involved in the design and delivery of climate-health education in schools delivering these programs. Course coordinators and program conveners most likely to provide climate-health offerings were identified from institution website listings of MPH and medicine degree programs, or through personal networks. Contact details were obtained from official lists (e.g. contact details on course curriculum websites), 48 eligible participants were identified, spanning 24 institutions, 40 schools and programs of public health (n=17) and/or medicine (n=23) (16, 17). Participants were recruited via email contact from the lead author with brief background information on the study, and an invitation to participate. If no response was received, one follow up email was sent three weeks after the original invitation. Participants were invited to take part in an online quantitative survey, self-administered online via the Qualtrics platform (Qualtrics, Provo, UT). Upon survey completion participants were invited to take part in an additional exploratory qualitative interview on the same topic. Survey and semi-structured interviews were conducted from February 3, 2021 to April 15, 2021. The ethical aspects of this research were approved by the Australian National University Ethics Committee (protocol number 681/2020).

All participation was voluntary, and participants did not receive financial compensation. Unless participants self-identified to do a semi-structured interview, names were not retained, and email addresses were not linked with respondents. It was indicated in the survey instrument that the results of the survey would be aggregated before anonymous results were reported more widely.

## Quantitative survey

### Measures

The online quantitative survey was designed and developed by two researchers (AL, CS) with experience teaching environmental and climate change to postgraduate public health and medical school students at the Australian National University. The survey included 18 questions modified from the Columbia Mailman School of Public Health study (14) (for a full question list, see the supplementary material). One question pertained to the institution and the academic role of the participant completing the survey. Six questions related to the participants’ experience and length of time teaching and whether they taught into a standalone/integrated public health or medical program. The other 12 questions covered curriculum design, teaching methods, and delivery of the climate health educational activities. Response to some questions were conditional (based on an answer to a previous question); where this occurs, the number of participants responding to the specific conditional question will be reported.

### Analysis

Due to the exploratory nature of this study, no statistical tests were performed. Instead, data were analysed using descriptive statistics. Open-ended questions were analysed by thematic analysis derived from word frequencies. The team characterised responses to the online survey and interviews relative to the sampling frame to check for nonresponse bias.

## Semi-structured interviews

### Measures

This was an explorative qualitative study designed to gain a deeper understanding of the quantitative findings. The interview questions were based on a top-down (theoretical) approach using findings from current research literature and a bottom-up (inductive) approach consulting with participants’ regarding their experiences of teaching in this area.

### Procedure

Quantitative survey participants who self-identified to be part of a follow-up interview were contacted via email. Subsequently, all interviews were conducted over Zoom by the first author, audio recorded, and subsequently transcribed prior to analysis using otter.io service.

### Analysis

Qualitative interview content was analysed via a modified grounded theory approach, where two coders (AL, EW) independently reviewed interviews and reached consensus to identify themes, and then independently coded interview content according to those themes. It was expected that saturation (i.e. no new insights obtained in the interviews), would be reached within twelve interviews if the purposive samples were relatively homogenous. To identify if saturation was archived, the approach outlined in Hennink et al. (18) was taken. As the authors coded issues/themes, the point at which each additional interview was not raising additional themes was noted (code saturation), and the point at which new insights within those codes was also noted (meaning saturation).

## Results

### Participant characteristics

#### Quantitative survey

Of the 48 participants invited to take part in the quantitative survey, 21 responded (43.7%) of which 16 were from Australia, two were from New Zealand and the remaining three participants chose not to report their institution. There was an approximately equal response rate for academics convening into public health (*n*=8) and medical (*n*=9) programmes, with two participants indicating Planetary Health and Health Sciences, and two participants indicating they did not directly convene courses.

Participants spanned a range of levels of seniority, with 23.8% (*n*=5) being professors, 23.8% (*n*=5) being associate professors, 14.3% (*n*=3) reporting holding PhDs, 14.35% (*n*=3) reporting senior lectureship, and one each of director and senior fellow (three participants did not specify their academic titles). The majority of the participants had taught into their programs for some time, with 52.4% (*n*=11) having taught for over 5 years, 38.1% (*n*=8) for between 2-5 years, and 9.5%(*n*=2) for less than two years. Over half (61.9%, *n*=13) of survey participants taught into postgraduate courses with 19% (*n*=4) teaching into both and 19.1% (*n*=4) teaching into undergraduate courses. Most educators rated themselves as moderately (33.3%, *n*=7), very (28.5%, *n*=6) and extremely knowledgeable (23.8%, *n*=5) regarding the association between climate change and health impacts. Three participants indicated they were slightly knowledgeable, or not very knowledgeable at all.

#### Qualitative interviews

A total of 10 people volunteered for a further face-to-face interview (4.8%) of up to 35 minutes long. Five participants lectured into Public Health programs and five into Medical Degree programmes. Participants represented a wide geographic region (two from New Zealand, three from Queensland and New South Wales each and one each from Western Australia and the Australian Capital Territory). Most respondents were at the Associate Professor or Professor level of seniority.

#### Nonresponse bias

Respondent characteristics for both online survey and qualitative interviews approximately equally represented public health and medical programmes. The original sampling frame consisted of 24 institutions (23 in Australia, one in New Zealand). Survey respondents represented 12 institutions, located in New Zealand, and in Australia in New South Wales, Queensland, Western Australia, and the Australian Capital Territory. Qualitative interviews covered the same geographical region. Accordingly, the sample does not represent perspectives from institutions in South Australia, Tasmania, Victoria, or the Northern Territory. While the online survey represented all levels of seniority, the qualitative interviews were biased toward more senior academic positions, thus views of postdoctoral early or mid-career researchers are under-represented.

### Quantitative survey results

#### Climate and environmental change content context and importance

Of the courses that participants convened or taught into, climate and environmental change was most often integrated into a wider public health or medical degree program (57.14%, *n*=12), rather than being a separate standalone course (23.8%, *n*=5), or having both integrated and standalone components (0.09%, *n*=2). Two participants did not specify course integration. Reflecting on the proportion of the total public health or medical degree program, all but one participant indicated that less than 5% of the degree lecture and assessment content focussed on climate change and health (90.0% of the 11 participants who completed this question). Figure 1 summarises brief open-ended reflections on why respondents think it is important for public health/medical students to learn about climate change. Emergent themes were that climate change was a health emergency and learning about extremes, adaptation and mitigation option and the role of social determinants of health in community were important. Academics also felt that students in public health and medical programmes had an important role to play as advocates and respected members of the community in voicing the importance of climate change as a challenge for health systems.

**Figure 1.**
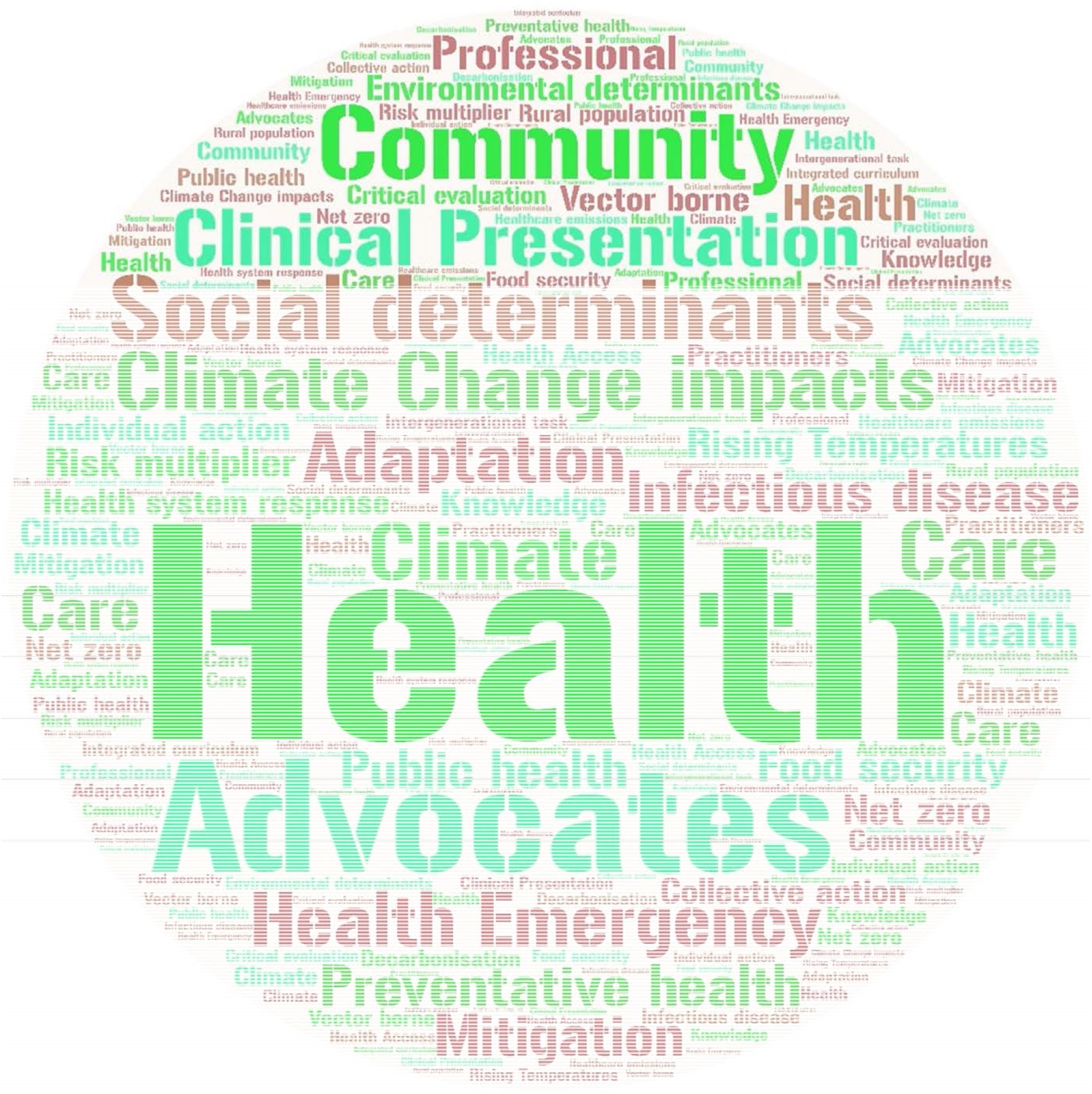
Word cloud from open-ended responses reflection on why it is important for public health/medical students to learn about climate change

#### Curriculum design and delivery

Reflections on the extent to which specific topics were covered in participant’s climate-health curricula are summarised in Figure 2. Notably, across all but one topic, participants rated coverage between “far too little” and “neither too little nor too much”. Only one topic, “health effects of environmental change” had any ratings indicating too much coverage, with one participant indicating this topic was covered “slightly too much”.

**Figure 2.**
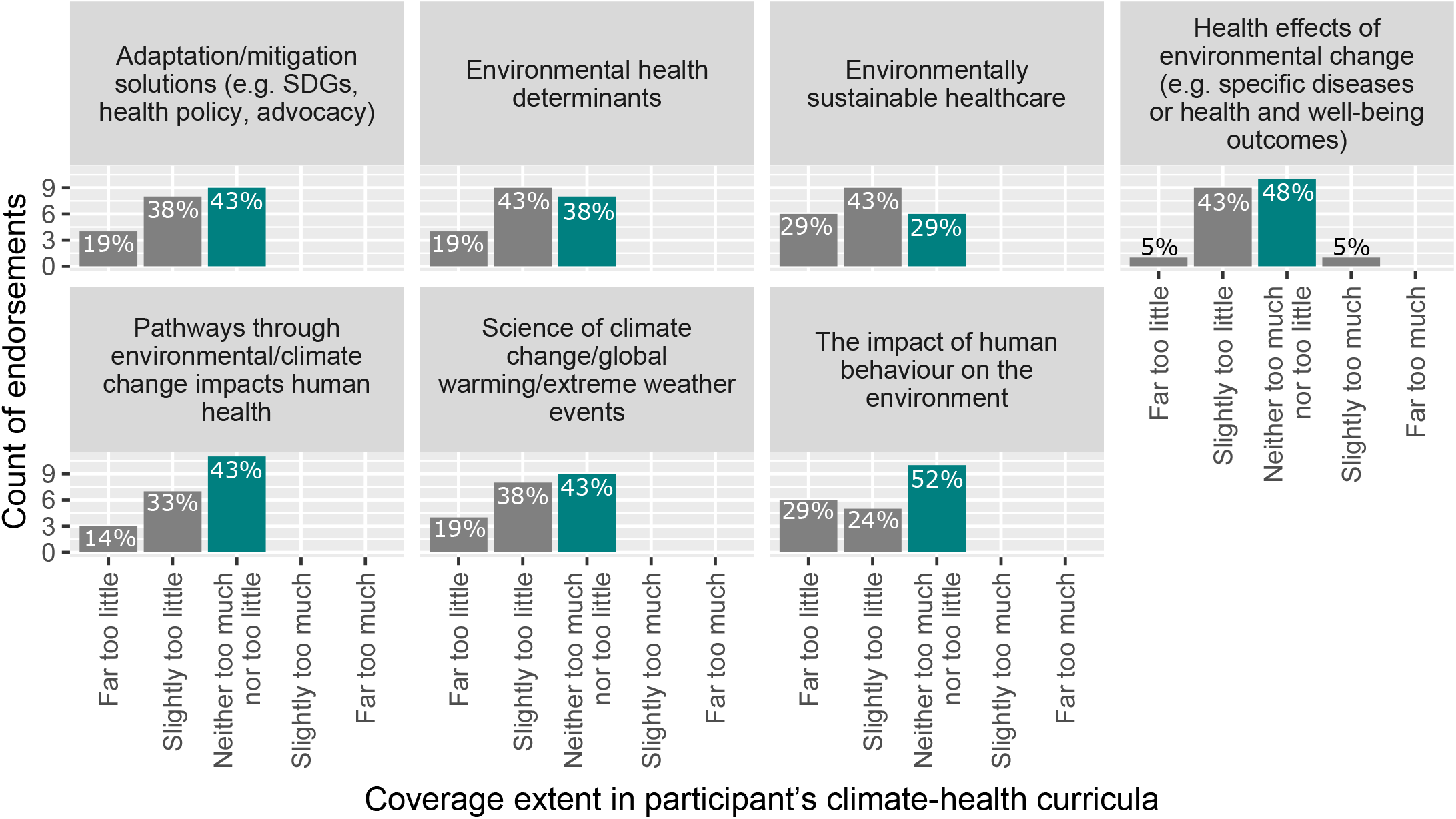
Summary of curriculum structure coverage in participant’s climate-health curricula

Two thirds of participants (66.7%, *n*=14) used more than one method to assess climate-health knowledge. The most common method was quizzes (used by 52%, *n*=11), followed by essays (used by 48%, *n*=10), exams (used by 38%, *n*=8), critical appraisal of journal articles (used by 19%, *n*=4), peer teaching (used by 14%, *n*=3) and capstone work (used by 14%, *n*=3), with only 5% (n=1) of participants reporting using placements or internships outside of their universities.

Subject matter was updated largely through reliance on subject matter experts brought in to give lectures (endorsed by 71.4%, *n*=15 participants), the scholarly literature (endorsed by 47.6%, *n*=10) and consultation with academic colleagues (endorsed by 52.4%, *n*=11 participants). As can be seen from Figure 4, the most common expert role for informing course content/guest lecturing was epidemiologists, followed by clinicians and social scientists, and environmental scientists. In relation to future offerings, nine participants (42.8%) indicated there was nothing new being considered. Five responses (23.8%) mentioned a standalone elective course being considered, four (19.1%) mentioned the possibility of a session in a required course while three responses (19%) mentioned climate-health doctoral degrees being considered. One participant (4.7%) mentioned existing partnerships with other academic institutions and non-academic institutions (business, government, NGO) in relation to course structure, two (9.5%) participants indicated partnerships with academic institutions and four participants mentioned existing partnerships with non-government institutions.

**Figure 3.**
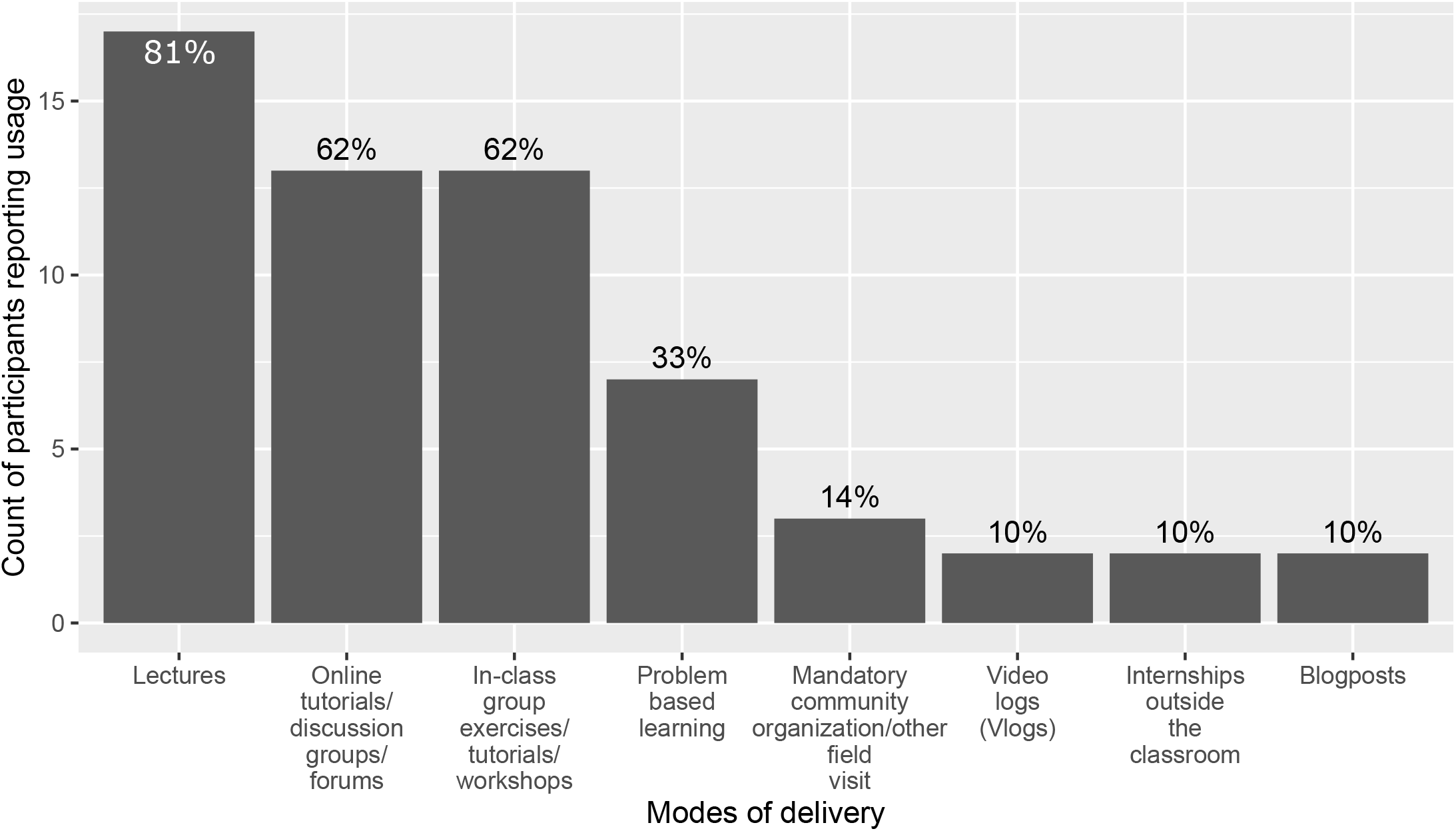
Summary of modes of information delivery

**Figure 4.**
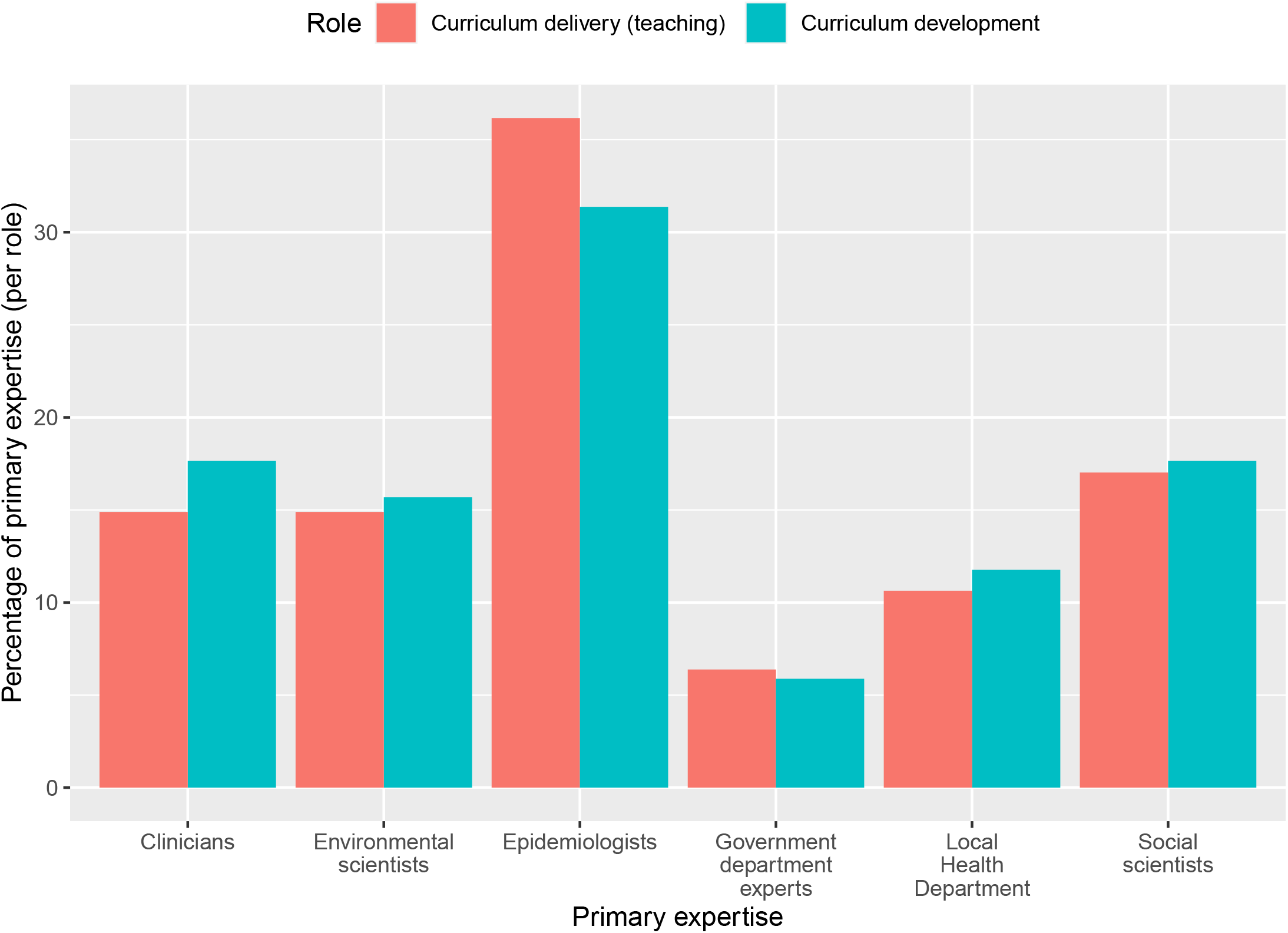
Expert roles in development and teaching of climate-health offerings

### Qualitative interview results

#### Themes identified

Four overarching themes in relation to teaching climate and environmental change in public health and medical curricula were identified:

1. Indigenous perspectives
2. Barriers and facilitators
3. Considerations in delivery
4. Future areas

Code saturation (the point at which additional interviews ceased to indicate the presence of new themes) was reached at approximately seven interviews. Meaning saturation (the point at which each theme is clearly understood) varied by overarching theme: saturation was reached for themes (1) (2) and (3), but heterogeneity in theme (4) indicates additional interviews may have revealed more information. These four themes were embedded within the motivation and pathway that led interviewees into teaching in this area, which will be summarised first to provide the context from where the themes emerge.

#### Motivation and pathways to teaching climate and environmental change

The interviews raised two common threads which framed the importance of climate change teaching in public health and medical curricula. First, almost half the interviewees raised the large carbon footprint of healthcare as an important factor driving their motivation to educate students about the impacts of climate change, believing the students would have an obligation to help reduce this in their future careers as doctors and public health practitioners.

> “*Health, like all sectors has to decarbonize……If global healthcare were a country, it would rank number five in the world in terms of the amount the size of its footprint*.*”*
>
> *“[we should] help professional graduates to be able to personally and professionally be able to reduce the footprint. So I feel very strongly about that as well*.*”*

The second common thread was the role of the interconnectedness between the global environment, and social drivers that the future health workforce should be aware of and consider when making personal and professional decisions.

> *“The thing I often try and get across if I’m teaching about climate change is that climate change is just one example of a global environmental trends, and that it’s very much interlinked and interlocks with diversity loss, poverty and inequality*.*”*

These threads informed participant reflections on their pathway into teaching in this area. This was often a process driven by individuals passionate about the topic, identifying a gap in the curriculum, and either developing their own curricula or advocating for the inclusion of an environment and/or climate change expert in the course delivery team. In a third of participants, this pathway began with sessional or guest lecturing into courses run by others.

> *“As a guest lecture [during PhD], to talk about climate change research methods in I think, undergraduate course. Then, when I moved to [city removed], as an academic staff, I discovered the gap in the curriculum in the teaching in public health, because there was zero content on climate change and health*.*”*
>
> *“The undergrad teaching, I more or less developed the curriculum on my own*.*”*

When reflecting on the teamwork underlying the delivery of environmental and climate change content, participants indicated working with an eclectic range of expertise, including policy consultancy, clinical practice, planetary health, field epidemiology, bioethics, business, and veterinary sciences.

#### Theme 1: Indigenous perspectives

An Indigenous perspective was universally seen as valuable in the design and delivery of environment, climate change and planetary health material.

Interviewees talked about the strong traditional connection to land and water as being a critical consideration in understanding environmental and climate change.

> *“We ignore their knowledge at our peril*.*”*

Indigenous perspectives were seen as particularly important within the realm of health, due to existing health inequities and the differential impact that climate change may have on their health in the future. Several discussed an understanding that different Indigenous peoples may have different experiences of loss as a result of losing their environments due to climate change, and expressed the importance that future doctors and public health practitioners are sensitive to this and how online teaching methods may not be the most appropriate medium to deliver some content.

> *“The gap already exists in many health issues there. And climate is just another layer. And I think that the health equity inequity between Indigenous and other communities will be exaggerated by climate change, definitely*.*”*
>
> *“There are certainly Aboriginal Health curricula that we can do on the online environment. But this sense of Country and sense of loss of Country, that’s gonna need a lot of thought before we include that into this particular medium*.*”*

All the interviewees agreed on the need for Indigenous-led input into course development and design. This was achieved through inviting community leaders, academic colleagues, State Departments with expertise in the area and long-standing practitioners in communities. Integration of these perspectives to some degree was generally possible due to strong connections within departments and with external stakeholders.

> *“And Aboriginal environmental health is part of the [State Government] plan I work within. So I call on my colleagues there to come in and talk about the issues that that we have in Aboriginal communities*.*”*

Some respondents indicated that they, regretfully, did not have the resources to properly engage with and reflect an Indigenous perspective in their course design or delivery, and did not feel it was appropriate to deliver such content matter themselves.

> *“You know, people like me who come from colonial roots from Europeans who have come here, don’t have those same connections, how could we [teach], you know, when you don’t have that long connection to the land*.*”*

#### Theme 2: Barriers and facilitators

The most salient barriers and facilitators to the inclusion and delivery of environment and climate change into medical and public health curricula were time and resources, and appetite from decision makers, contributors, and students.

#### Time, resources and an already full curriculum

The most common barrier - endorsed by all interviewees - was the issue of limited time and resources. This reflects the fact that environment and climate change is one of many competing priorities within the full curricula being delivered.

> *“I would like to say we could do more, but I actually don’t think we can do more. In addition, something would have to come out. No one ever wants to give anything up in the curriculum, ever. So I think there’s this constant pressure on it*.*”*
>
> *“It’s not lack of interest. It’s just lack of time*.*”*

#### Appetite from decision makers (politics, institutions, culture)

Decision makers, and the role they play in how curriculum is organised and taught, were raised as a vital component of successfully integrating environment and climate change material into courses and programs. All interviewees talked the role of top down support as essential to bringing about change in the curriculum. Two interviewees mentioned national politics as a barrier to getting traction in this area.

> *“It’s all political*.*”*

Institutional support and support from key individuals within institutions, such as heads of school and members of committees who drive curricula, was also seen as important in allowing more climate and environmental change to be embedded into the curricula. One factor underlying institutional support raised was awareness of the importance of environment and climate change for health.

*“There’s institutional barriers about who [is] allowed time in the curriculum*.*”*

> *“So the first thing you got to do is educate the faculty about the importance of these issues. Because otherwise they don’t necessarily, you know, they’re just not aware of it*.*[…] A lot of that teaching is really done by more senior academics, or at least if not done by them, supervised by them. They are the ones who sit on the committees, the curriculum committee, for example, that so they’re making decisions about what will and won’t be given space and the curriculum*.*”*

A barrier to successfully raising awareness and convincing institutional decision-makers of the importance of environment and climate change for health, raised by two interviewees, was the historically conservative nature of medicine as a profession, and conservative views held by more senior practitioners and educators.

> *“It’s extremely conservative profession. And it is slow to change and slow to adapt to new things. And so I think it’s been slow to adapt very, very slow to, to start work on dealing with Climate change*.*”*

On the flipside of these barriers, the role of key individuals and high-level support in giving prominence to climate/environmental change in the curricula was raised by most interviewees as a critical factor driving its inclusion.

> *“The only way to do it is to have champions*.*”*
>
> *“I think, for me, it’s lucky because the Head of School is very supportive in developing this new curriculum*.*”*

Reflecting the interviewee’s personal motivations and experiences leading to their teaching environment and climate change, there was an acknowledgement that bottom-up momentum was vital to meet any opportunities arising from top-down support.

> *“At the minute, it’s from the ground upwards, because I don’t see a lot coming from this side, even though even though organizations like the AMA, the Australian Medical Association have declared a climate emergency*.*”*

#### Appetite from contributors

Academics also raised the role of contributors (lecturers, clinicians) as an important factor in making sure that climate and environmental change and planetary health were prominent in the curriculum. The need for contributions could be difficult due to siloed views, large time commitments to projects and the onus on individual course conveners and coordinators to drive the environmental health agenda forward.

> *“They sit in their silos and they don’t get out and they don’t break down those cross organisational borders. They’re really acutely aware that they’ve got these social silos, but it also goes up to organisational silos as well*.*”*
>
> *“All the supervisors [are] working gratis, this has been an enormous amount of time guiding the students meeting with them last year, largely online. […] Within our population medicine theme, and had it not been for us pushing it, it would have really struggled to get a place in the curriculum*.*”*

However, positive reactions from individuals who could contribute to the course were seen as highly beneficial.

> *“And I actually grabbed all the opportunities to talk about climate change and health with my colleagues here in the school. […] The colleagues are really appreciating the opportunities to engage with this topic. So then I think you need to be proactive*.*”*

#### Appetite from students (willingness to engage/ask for the course)

Most academics mentioned the importance of student engagement in the topic as a key driver for making it fit into the curriculum. A common thread within these discussions was that students were often reluctant to engage at first, but their views became substantially more positive upon understanding the importance of environment and climate change for health.

> *“So we have to craft our way in communication in you know, getting students to understand the issue first. So they would be interested to know more to understand why we have to discuss this in the classroom. And I think that’s very challenging actually. Once you get people engaged and involved, and they are, I think many of them are passionate about what they can do to solve the problem*.*”*

#### Theme 3: Considerations in delivery

Almost all of the interviewees recognised the importance of delivering information in a manner that students found engaging and thought provoking. One major component of achieving this was choosing the correct lecturers or being passionate themselves. The other was through avoiding delivery formats that were inappropriate for the students (especially those with clinical and caring responsibilities) and favouring active discussions with positive framing.

> *“It really boils down to being your how engaged you are with the subject matter, I think, and how enthusiastic you are about the subject matter. It comes through it in your teaching, and therefore, the students get more out of it*.*”*
>
> *“Students hate group work. And I hate it with a passion*.*”*
>
> *“I think most students are keen on solutions-based discussion rather than [focussing on] the problem*.*”*

Forms of delivery raised as successful examples of promoting discussion included direct interaction with guest experts, online discussion forums, discussion of contemporary evidence (ideally, recent manuscripts authored by the lecturers themselves), and assessment formats focussing on critical thinking. Many interviewees indicated that their choice of delivery format, and what content was taught, was driven by considering the future trajectories of students.

> *“We’ll come back to what type of skills are we trying to teach the people that were taught?*
>
> *You know, that were one of the one of the future roles and what are we trying to teach them?”*

The specific environmental and climate content taught was motivated to some degree by the interviewee’s own expertise, guest lecturer availability, and availability of contemporary scholarly publications (many authored by the interviewees themselves) that could serve as a basis for discussion. One interviewee noted clinical accreditation standards tended to guide learning objectives, and consequently the nature, space, and content afforded to topics such as environment and climate change. Two interviewees raised the importance of the Australian Medical Association, both for setting this agenda, and as a source of topics for delivery.

> *“They [the Australian Medical Association] want to see the health sectors emissions cut by 80% by 2030. So we need to be doing this*.*”*

At the time of interview, all interviewees had experienced an obligatory shift to online course delivery for at least one semester due to the COVID-19 pandemic. Accordingly, the bulk of reflections on course delivery focussed on the transition from previously face-to-face to online teaching via platforms such as zoom and skype. Interviewees varied in their prior experience of online teaching, from no experience through to having provided multiple years of online delivery. For delivering content to students with caring responsibilities, jobs, and especially those with clinical responsibilities, the transition to online was seen as beneficial. Yet, some interviewees noted students raised concerns that online delivery decreases engagement and interactivity, particularly among students.

> *“[this change] demonstrated fairly clearly to us that we needed to move to meet the demands of the students that we have. the feedback that we’ve had is that the students really appreciate the flexibility […]with that cohort being, you know, medical practice, practitioners, nurses, etc, for it for GP doctor to take two weeks out of his practice, to come and do a course is almost impossible. It costs him 1000s and 1000s. of dollars*.*”*
>
> *“I think some of the students did express their concerns about lacking of engagement in the online community*.*”*

#### Theme 4: Future areas

Moving on from reflections on the history and current practice in teaching environment and climate change in public health and medical curricula, interviewees were asked to reflect on the future of their courses. Approximately half of interviewees reported an intention to expand cross-disciplinarity and education outside the classroom. Key areas highlighted for this expansion were policy, and local community engagement. This was discussed in terms of expanding the topics within existing delivery and seeking guest lecturers working in policy or embedded in climate change affected communities. Two interviewees further emphasised their desire to engage more with communities in future, indicating the prospect of more sustainably involving community input into their courses, and seeking to expand the impact of their teaching to the community level.

> *“Making it a more holistic programme where it becomes part of the thinking of the future medical doctors to think about these issues when they’re thinking about clinical issues*.*”*
>
> *“Population capacity building, community education, I think, particularly this age of disinformation, got a big challenge to not just try and educate people within the universities, but also to reach out and try to try to interest and educate whole communities*.*”*

Several of the interviewees noted that they intended to refresh their course content. This was typically related to the recognition that more recent topics, case studies, and scholarly manuscripts would enhance student engagement. Other motivations included creating a better fit at the program level by adjusting to changes in other courses and pivoting to reflect the current research of themselves and colleagues contributing to the course. One interviewee noted that ensuring the course content was contemporary was especially important for the student’s future practice.

> “*And the diversity of patients they’re going to see, for a specific condition, be it by age, group pain, by gender, by ethnicity… So that’s where there will be further fine tuning*.*”*

Alongside expanding content, interviewees discussed opportunities for expanding the forms of delivery they could offer. While many touched on the possibilities that online delivery facilitated, their discussion generally did not extend to concrete intentions to experiment with online formats in ways that would change the course structure as a whole (e.g. shifting a semester-long course to an intensive or year-long online-only format). One interviewee raised their intention to meet calls for the micro accreditation (aka micro credentials) format from organisations such as the Public Health Association of Australia but did not pursue this due to lack of support from their institution’s leadership.

In terms of the future pedagogy of their teaching, the predominant thread across interviews was creating a possibility for students to engage more with researchers. One interviewee invoked the “capstone” paradigm as a method of ensuring students could make the most of a research-intensive experience.

> *“I would like to develop more than we do at the moment opportunities for being active in the student group, or other groups doing research in this area. So in the new curriculum, I am pushing for capstone experiences. Which could be a research project*.*”*

Another source of pedagogical change was practice in related fields.

> *“I like going back to disaster management structures in that multifaceted approach very much. Because it actually works quite well. You know, people out of their comfort zone with a common purpose. Yeah. And maybe we can take that forward into our teaching in public health in different ways as well*.*”*

## Discussion

Broadly consistent with global studies, our findings indicate that climate change is not consistently integrated into health professional training (14, 19, 20). Our results offer a more nuanced assessment of the barriers and opportunities for making climate change more central in medical and public health curricula across tertiary institutions in Australia and New Zealand. Notably, the limited integration of Indigenous-led teaching and learning into curriculum design and delivery was apparent in both public health and medical programs, despite universal recognition of the important role of this knowledge when teaching students about the interconnections between our environment and health. Strengths of our study include an in-depth quantitative and qualitative assessment of medical and public health programs in the Southern Hemisphere. While large scale surveys of health professionals and the role of climate change in health curricula have recently been carried out (14, 21), these studies focus on institutions in the northern hemisphere, do not include a qualitative component or perspectives from both public health and medical programmes. Although climate change has been an issue for decades, it appears that Universities in Australia and New Zealand are still struggling to integrate this into training programs for our future health workforce.

We provide a geographically based starting point to strengthen the environment-health curriculum at the graduate level for Australia and New Zealand. However, non-response bias cannot be excluded due to the modest response rate (43.5%). Institutions that are progressively engaging with curriculum development in the area of climate-health education are likely to be over-represented as academics will have been more likely to participate in the study. The integrated nature of medical programs made it difficult to identify the most suitable academic from institutional websites to invite to participate in the study, relying in part on the authors’ knowledge through professional networks. Future studies could include academics teaching into these programs who are not engaged in teaching about climate change and planetary health as well as a broader range of health professions programs such as nursing. This is likely to provide further important insights into the challenges of teaching in this area. Our study also did not include organisational leaders, an important subgroup identified by interviewees to engage with.

Our findings suggest that there are existing facilitators within public health and medical training programmes that can be strengthened. However broader engagement and uptake of the value of teaching these students about climate change will move slowly without explicit strategies to further increase student and community engagement with the issues, address institutional barriers, and build leadership and capacity in the next generation of teachers in this area (22).

The lack of visible Indigenous-led education in this area represents a significant missed opportunity for health leadership. Given the strength of Indigenous knowledge in this area and their comprehensive understanding of the connections between our environment and health, there exists an enormous (largely untapped) opportunity to embed a strengths-based approach to Indigenous environmental health, further support the growth of the Indigenous health workforce and enhance the cultural competence of our non-Indigenous future practitioners.

### Targeting institutional barriers and building academic leadership

Our findings indicate that workforce capacity, limited leadership and formal training opportunities in interdisciplinary research and teaching, and entrenched views of climate change in established faculty were persistent barriers across public health and medical curricula within institutions. While majority of our survey participants had taught in this area for over five years (52.4%), connections with other academic institutions, non-academic organisations and non-government institutions for the purposes of teaching were low. The role of institutional leadership or having high-level “champions” was highlighted by majority of the interviewees as a key facilitator to achieving an integrated and visible climate change and health curriculum in public health and medical programs. Well-resourced communities of practice that include links with community and government could help improve interdisciplinary teaching of such “wicked problems” by enabling staff leadership and institutional scholarship (23). Interviewees highlighted the use of capstone experiences and micro-accreditation to further embed multidisciplinary perspectives into teaching programs. All interviewees reiterated the importance of providing a holistic understanding of environmental and social issues on health (24), both as a key driver of their current content and an area of future focus. For most programmes, epidemiologists played a major role in designing and delivering the content. What is needed is more structural support to develop the development of academic teams teaching this content. This will not diminish the important role of epidemiologists, but rather recognise the importance of diverse disciplines contributing to the design and delivery of a holistic programme.

Interviewees also raised the point that the flexibility of online learning due to the pandemic resulted in an exponential increase in enrolments by practitioners. This offers another opportunity to re-think course design as pre-recorded content may help alleviate some of the time constraints felt by all educators and allow experts from external organisations to have a bigger role in teaching through video-links. This shift is also an opportunity to further embrace modern blended learning approaches such as online and face-to-face tutorials/workshops, discussion boards and forums, blogposts and video logs, which are currently used infrequently, as indicated by our survey results. The development of the *MJA-Lancet* Countdown indicator to track Australia’s medical schools in preparing future doctors to deal with the current and future environmental threats and work in an environmentally sustainable health care system alongside other Australian and New Zealand designed indicators (see Section below) will set a benchmark against which these models of delivery can be measured (25, 26). Building an effective community of practice around effective interdisciplinary teaching and learning will require targeted investment and resources.

### External influences on curriculum

The external regulatory framework was raised as a key factor in driving change in curricula. For example, the Association for Medical Education in Europe (AMEE) Consensus Statement on planetary health and education for sustainable healthcare (10) recommends an eco-ethical leadership approach to achieve the education for sustainable health care vision: “integrated around environmental and ecological sustainability, values, collaboration, justice, advocacy, and activism designed to address issues in complex systems” and incorporating diverse cultural views. The values exhibited by institutions, such as the ANU Below Zero initiative (27) and the University of Melbourne Sustainability Charter (28) that make commitments to sustainability in their operations, research and teaching are vital to enacting such a vision. The development of recommended targets and indicators for use by health professional educators, institutions, and regulators to implement and evaluate education for sustainable health care (10, 29) is timely, given the AMC Review of Accreditation for Primary Medical Programs that is currently in progress (30). The Council of Public Health Institutions in Australia and New Zealand (CAPHIA) has included understanding the theory of climate change and its implications for sustainable development as a core competency underpinning health protection curricula (17).

The political environment was also raised by interviewees as an important challenge to the delivery of climate change in the health curriculum. One of the recent challenges was the last US president pulling out of multinational initiatives like the Paris Agreement (31, 32). One interviewee mentioned that the change in government in the US in 2020 would likely have positive flow on effects for teaching climate change, not only in the US but also in Australia. Conversely, such obstruction has also seen other countries step up to make climate change a focus in university teaching. In a recent study of academics teaching climate change in China, a new strategy for climate change inclusion in higher education institutions was adopted in response to the US rejection of the Paris Climate Agreement (33). □These external influences impact on the ability and capacity of public health practitioners and academics to create an integrated climate-health curriculum (34). A counterpoint to this is the important role of advocacy raised by all our interviewees. Advocacy organisations like the “Climate and Health Alliance” and “Doctors for the Environment” were seen as instrumental to help catalyse public action to support more climate change action to protect health. The climate strikes were also highlighted as an event that galvanised student and academic action around climate change and health. Partnering with advocacy organisations to deliver content and leveraging global events such as the climate strike to publicise the research and trigger further action may be a future opportunity to elevate the need for a comprehensive, contemporary climate-health curricula.

### Indigenous-led environmental health curricula

Climate change adaptation and resilience will need to be based around connection to Country and cultural values. The differential impact of climate change on the health of Indigenous populations was seen as a particularly important aspect that needed to be embedded into the curricula and be Indigenous led. This is consistent with the finding of Cameron (35) who found that the well-intentioned aim to improve the lives of Indigenous northerners in the Canadian Arctic was often rooted in climate change adaptation frameworks, knowledge and practices that perpetuated colonial assumptions. There is clear evidence that Indigenous health professionals and practitioners deliver improved healthcare outcomes for Indigenous patients and communities (36, 37). In addition, non-Indigenous staff deliver better outcomes when working with Indigenous staff and these partnerships are critical to the provision of culturally safe care and to reduce institutional racism (38, 39). Similarly, Indigenous-led climate-health education incorporating Indigenous knowledge systems is crucial to enhancing the social and environmental accountability of medical and MPH progams (40). The potential for enhanced adaptation solutions and better health outcomes in the face of climate change cannot be fully realised without an overhaul of the currently inadequate structural support to grow Indigenous environmental health leadership.

## Conclusion

Climate change is a major threat to human health and survival. Climate solutions represent one of the biggest opportunities to advance health, well-being and reduce inequities. Universities have a key responsibility in leading the development of these solutions for society. This first mixed-methods assessment of climate-health education across tertiary institutions in Australia and New Zealand shows that currently, progress in incorporating sustainable health care, climate change, and planetary health into curricula rests with motivated individuals. For most educators this action is missing a systematic approach without universal support and strategies within and outside institutions to enable change. The ad-hoc Indigenous-led environmental health curricula is a missed opportunity to deliver culturally informed climate-health content. We recommend that building leadership through stronger partnerships with policymakers, community stakeholders and advocacy organisations will help reshape the core curricula in medical and public health schools so that it is contemporary and coherent amid an increasing climate health risk. In addition, curricula review incorporating targets, changes to accreditation standards and indicators to track progress will provide much needed structural support and impetus to enable systemic change within institutions. Now is the time to reframe how climate change and broader environmental and related social issues are integrated into the curriculum to best equip our future health workforce.

## Data Availability

This was a mixed methods study involving a survey and qualitative interviews. Data has been used in the manuscript and aggregate data are available upon request

---

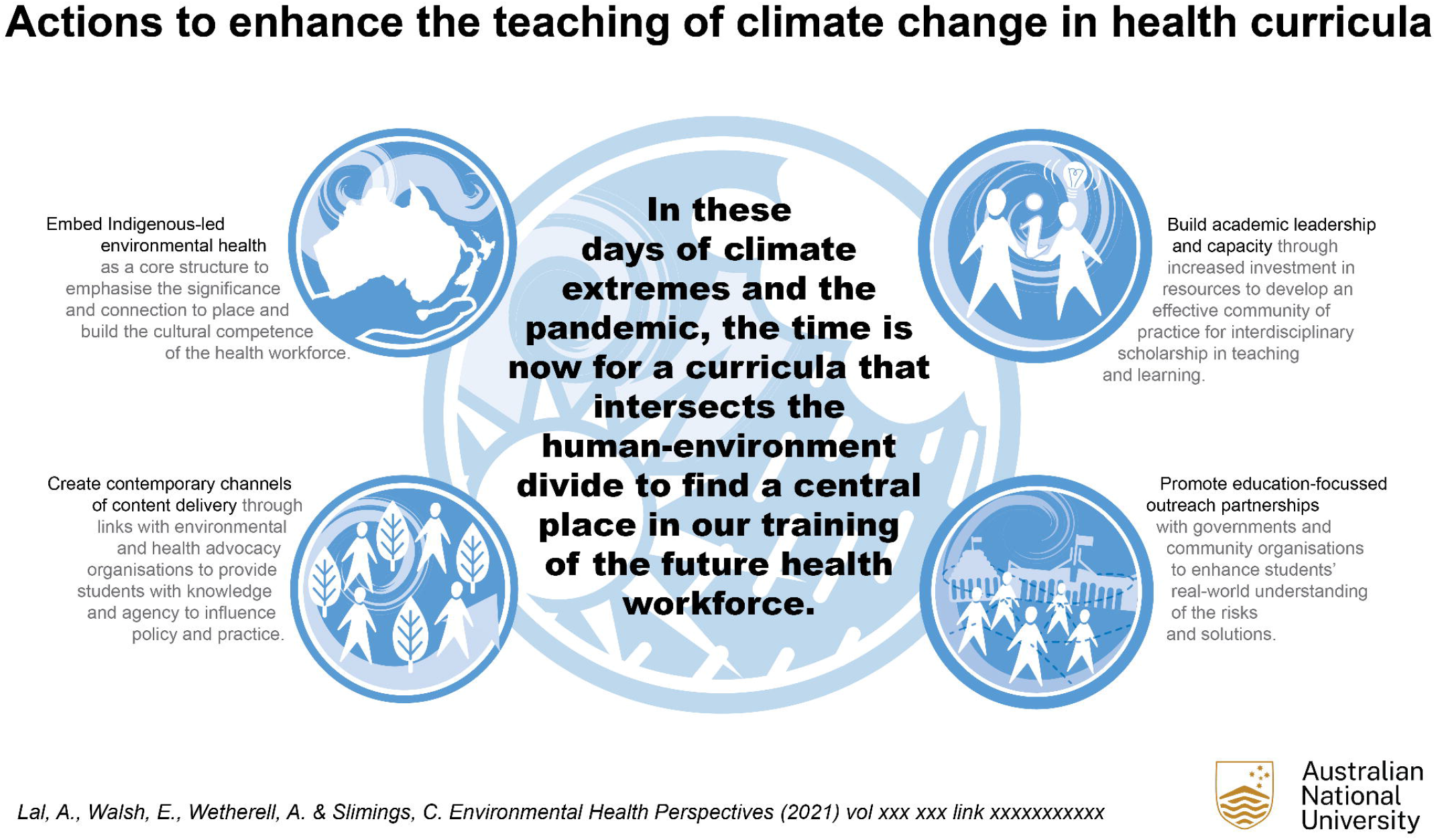

