## Supplemental file for "Climate change education in public health and medical curricula in Australian and New Zealand Universities: a mixed methods study of barriers and areas for further action"

Supplemental Material

Study questionnaire

**Background Information about your Institution and teaching experience**

1. Is the program you teach into an undergraduate or postgraduate course?

- Undergraduate
- Postgraduate
- Other (please specify)

2. How long have you been teaching into the programs mentioned?

- Less than 2 years
- 2-5 years
- More than 5 years

3. Do you convene any programs related to Public Health or a Medical degree?

- Public health
- Medical degree
- Both
- Other (please specify)
- Do not convene

4. Is climate change integrated into the program you convene or a separate course within the program in your university?

- Integrated (please go to Q5)
- Separate stand-alone elective course for the Degree
- Separate stand-alone core course for the Degree
- Separately offered as a Masters/Graduate Certificate program
- Capstone or leadership program in Climate change and Health
- Other (please specify)

5. What percentage of the degree (approximately) focusses on climate change and health (percentage of lectures/assessments)?

- Less than 5%
- 5%-10%
- More than 10%
- More than 20%

**Teaching and Delivery**

6. Why do you think it is important for public health/medical students to learn about climate change? *Maximum 500 characters.*
7. Which staff are involved in the **teaching** of climate change- health curriculum in your school? *Please tick all that apply.*

- Clinicians
- Epidemiologists
- Environmental scientists
- Social scientists
- Government Agencies
- Local Health Department
- Others (please elaborate)

8. Which staff are involved in the **development** of climate change- health curriculum in your school? *Please tick all that apply.*

- Clinicians
- Epidemiologists
- Environmental scientists
- Social scientists
- Government Agencies
- Local Health Department
- Others (please elaborate)

9. To what extent are the following topics are covered in your climate-health curriculum? (Rated as *1 “far to little” through to 5 “far too much”)*
   - Science of climate change/global warming
   - The impact of human behaviour on the environment
   - Pathways through environmental/climate change impacts human health
   - Health effects of environmental change
   - Environmental health determinants
   - Environmentally sustainable healthcare
   - Adaptation/mitigation solutions (e.g. SDGs, health policy, advocacy)
10. What resources do you use to design and update curriculum/topics taught?
    - Bring in subject matter experts and ask them to provide updated content
    - Rely on scholarly literature to update the curriculum yourself
    - Consultation with academic colleagues
    - Topic specific feedback from students
    - Local/Regional community of practice with shared resources (e.g. case studies and tutorial materials)
11. What are the main learning outcomes of your overall climate-health curriculum? *Maximum 500 characters.*
12. What teaching methods are used to deliver climate-health content to students? (Please select all that apply)

- Laboratory based exercises/practicals
- Lectures
- In-class group exercises/tutorials/workshops
- Online tutorials/discussion groups/forums
- Internships outside the classroom
- Video logs (Vlogs)
- Blogposts
- Mandatory community organization/other field based visit
- Problem based learning

13. What kind of case studies do you use in your teaching?

- Mainly international
- Local/National
- A mixture of national/international

14. Does your school currently have any education related partnerships on climate change and human health? (Please select all that apply)

- Yes, with another academic institution
- Yes, with a non-academic institution (business, government, NGO, etc.)
- Yes, with a funder
- No

15. How is climate-health knowledge assessed in your course? (Please select all that apply)

- Quizzes
- Exams
- Essays
- Critical appraisal of journal articles
- Capstone
- Peer teaching
- Placements or internships outside the University
- Other (please elaborate)

16. Are any climate-health offerings under discussion to add? (Please select all that apply)

- Session as part of non-required course
- Session as part of required core course
- Climate-health standalone elective course
- Climate-health standalone required course
- Climate-health masters or certificate program
- Climate-health doctoral degrees
- Climate-health post-doctoral positions
- Nothing being considered

17. How knowledgeable do you, as an educator feel about the association between climate change and health impacts?

- Very knowledgeable
- Moderately knowledgeable
- Not very knowledgeable
- Not knowledgeable at all

18. Please specify the name of your institution, your academic title, and role.
19. Would you consider a short semi-structured interview to follow this ( 30 minutes)

- Definitely yes
- Might or might not
- No
